# Anatomically-guided deconvolution of PET using directional total variation regularization

**DOI:** 10.1101/2023.04.23.23289004

**Authors:** Ashley G Gillman, Min Han Quah, Evangelos Papoutsellis, Kris Thielemans, Pierrick Bourgeat, Claire Delplancke, Jurgen Fripp, Edoardo Pasca, Jason A Dowling

## Abstract

Positron emission tomography (PET) provides quantitative functional imaging of biomarkers unavailable in other modalities, however, images are of relatively low resolution compared to modalities such as magnetic resonance imaging (MRI). A typical approach is to reconstruct to a higher resolution and regularize using a structural image, but there are practical limitations to this approach. Alternatively, post-reconstruction approaches involve image-based correction, but typically rely on a segmentation which may be difficult or even ambiguous to find, depending on the anatomical region or deformities.

Here, we perform super-resolution by utilising iterative deconvolution, regularized by minimizing shared directional total variation (dTV) with an anatomical MRI image. We present results on synthetic and clinical data. For the former, PET acquisitions were simulated using an analytic PET simulation. The Gaussian blurring model parameters for deconvolution were optimized on a simplistic phantom simulation with a total variation prior. This model was then applied to deconvolve realistic synthetic data using dTV, which was synthesized to include PET-unique lesions. The model was also applied to a single 18F-florbetaben study acquired over 10 minutes.

Gray matter-white matter contrast increased using dTV compared with baseline, however, where an accurate segmentation is available, traditional partial volume correction techniques are superior. Hence, dTV-regularised deconvolution can perform PVC and super-resolution in situations where a reliable segmentation cannot be achieved. With appropriate hyper-parameter selection, dTV deconvolution can preserve PET-unique features.

## 1 Introduction

Positron emission tomography (PET) is an important imaging modality capable of providing information regarding the functionality of an organ. This aids in the diagnosis and management of diseases such as cardiovascular diseases, oncology, and neurological disorders. However, the resolution is limited in comparison to other anatomical imaging modalities such as magnetic resonance imaging (MRI) and computed tomography (CT). Limited resolution can result in missing small clinically-relevant features such as lesions, inaccurate quantification of biomarkers, and inconsistent quantification of biomarkers between different hardware. This has led to a desire for PET super-resolution techniques, especially with increasing popularity of hybrid PET/MR scanners.

The partial volume effect (PVE) describes the problem that the intensity of a given voxel of an image represents an aggregation of a region of the object being imaged. One cause of PVE is the limited resolution of the image representation – practically this means that a given voxel represents a sum or average of the intensity within its volume. In PET, PVE is further complicated by the point-spread function (PSF), which is a model of the intensity dispersion of the imaging system and is generally the limiting factor of the effective image resolution, regardless of the reconstructed PET image voxel size. PSF can model the physical effects of photon non-collinearity, positron range and detector depth-of-interaction; hardware effects of detector size and geometry; and practical effects of reconstructed voxel size and axial/transverse mashing [1]. Although some of these phenomena are more naturally modelled in sinogram space, the PSF encompasses all effects after image reconstruction, and models them with either a spatially-invariant [2] or spatially varying kernel [3]–[7].

In this work, we use the term “PET super-resolution” to refer to techniques for resolving PET images beyond the physical resolution of the scanning system. It consists of both upsampling and partial volume correction (PVC). This means that the problem becomes ill-posed, and therefore it is necessary to incorporate prior information [8]. Often, the prior may be defined with respect to an anatomical image. Anatomical images are typically MR or CT images, although preferably MR due to the increased soft tissue contrast.

Traditional techniques for PVC rely on the PET image being segmented into regions-of-interest (ROIs). In neuroimaging, this means a tissue segmentation (grey and white matter and cerebrospinal fluid), or segmentation of lobes or gyri. The requirement for segmentation implies a dependency on the accuracy of the segmentation algorithm and can prove difficult or ambiguous, especially in the presence of structural abnormality. The first class of PVC correction are region-based PVC, wherein a crosstalk matrix is developed describing the mean observed values in each region as a weighted sum of the underlying true regional mean values, and inverting these to solve for the corrected regional means. The most simple of these approaches calculate a recovery coefficient for a single region, either ignoring [9] or including [10] spill-in from background regions; but other approaches model more complex mixing between all regions [11], [12]. Traditional voxel-based approaches extend these methods to be able to provide correction at the voxel level, but still consider the effects of spill-in and spill-out at the regional level and hence require tissue segmentation [13]–[17]. These methods include the region-based voxel-wise correction (RBV) [18], which applies a voxel-wise correction to ensure regional means are close to that predicted by a regional mean method, and the iterative Yang (IY) algorithm [19], which iteratively applies such corrections. Another class of approaches utilises deconvolution. Without anatomical image-based regularisation [20], these approaches are prone to amplifying noise and producing artefacts. A comprehensive review of traditional approaches is given by Erlandsson *et al*. [2].

An alternate approach is to reconstruct to a higher resolution, incorporating PVC into the reconstruction by including an appropriate prior. Commonly, a smoothing prior is designed to only operate over regions with local similarity in the anatomical image. The Bowsher prior [21] limits smoothing to a subset of most-similar voxels in the anatomical image; whereas other approaches use weighted smoothing based on local similarity [22], [23]. Another approach attempts to force gradient changes to be shared between the reconstructed PET and anatomical images by regularising a weighted version of total variation (TV), where weights are calculated from the anatomical images. In regular weighted TV [24], weights reduce the penalisation in regions with high gradient in the anatomical image, whereas in directional TV (dTV) [25], [26], vectorised weights further encourage local gradient (i.e., edges) to be parallel or antiparallel.

A recent class of techniques pose PVC as an image-to-image regression problem, which is solved using deep learning approaches [27]. These approaches are promising and offer qualitatively superb and very fast results but are not currently suitable for quantification as there are no guarantees regarding consistency with the measured data. Belthangady and Royer [28] describe this phenomenon in the context of fluorescence image reconstruction as problems due to hallucination, where statistically common but absent features are added or enhanced in an image; and problems due to generalisation, where data outside the training data manifold are poorly reconstructed.

The traditional techniques offer an advantage in that no modifications to validated reconstruction methods needs be performed, and the operations can be performed entirely in image space. However, the reconstruction approaches avoid the need for a segmentation, although they can be used if available. A final class of algorithms are those that perform deconvolution and utilise a structural prior that doesn’t require a segmentation. Bousse *et al*. [29] propose a deconvolution with a regulariser based on a Markov random field that is weighted to only smooth intra-regionally within a segmentation. Wang *et al*. [30] extended this technique to regions with similar PET and MR structure, avoiding the need for a segmentation. These approaches are akin to reconstruction approaches where the forward model is restricted to PSF modelling without projection, and the authors believe is the only published technique for segmentation-free PET deconvolution.

In this work, we extend upon the work of Wang *et al*. [30] and formulate PET reconstruction as a two-stage optimisation with a separate reconstruction. We then extend the previous approach and present the first dTV-regularised PET/MR deconvolution results on simulated and clinical data.

## 2 Methodology

Image reconstruction can be generally stated in the following generic formulation:

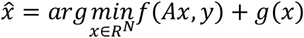

where *x* is a reconstruction image estimate with *N* voxels; 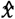is the estimated optimal reconstruction; *A* is the acquisition model, modelling the imaging system; *y* represents the measured data, typically a sinogram in PET; *f* is the data fidelity function, a similarity metric for data consistency; and *g* is a regularization function on the reconstructed image, encouraging the desired *a priori* properties. In PET imaging, the acquisition model is typically an affine operator that includes factors for the geometric projection from image to sinogram space, sensitivity, normalisation and attenuation and terms for random and scatter background. In addition, the acquisition model can include additional physical effects including motion-related resampling and, most relevantly, the PSF model of the scanner.

A typical approach to PET super-resolution is to increase the resolution of *x* beyond the physical limitations of the scanner. In this case, the reconstruction can become ill-defined without appropriate regularisation. In this work, we instead separate the reconstruction and super-resolution into sequentially nested optimization problems, such that:

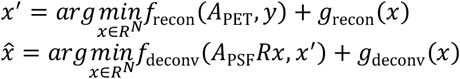

where *x*^,′^ at standard resolution, *A*PETPET acquisition system, *A*PSF is the PSF model in image space, *R* is an upsampling operator, and *f*recon, *f*deconv, *g*recon and *g*deconv are the data fidelity and regularisation functions for the separate reconstruction and deconvolution stages.

### 2.1 Anatomically-guided deconvolution using directional total variation

In this work, we used convolution with a multivariate Gaussian kernel, *K*, as the PSF model:

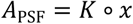

with the kernel defined at each voxel index, *i*, with location in physical space relative to the centre of the kernel, *l*i ∈ *R*^3^:

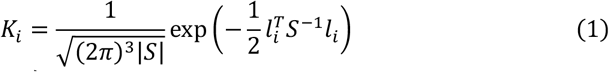

where *S* = diag(σXy, σXy, σz) is the covariance matrix, such that convolution in *K* separable in *x, y* and z and isotropic in *x* and *y*. σXy and σz are the standard deviations in x/y and z respectively, although we report these throughout the paper in units of FWHM.

We use the squared L2 norm for the deconvolution data fidelity term:

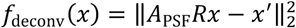

and dTV [25] for the deconvolution regularisation term, defined as:

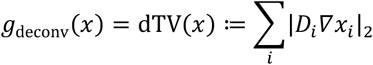

where *D* is a weighting vector field on the gradient in *x*, dependent on the normalised gradient of the anatomical image, *ξ*:

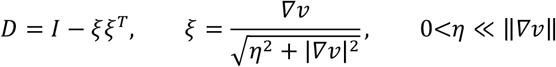

and where *η* is a small constant to avoid division by zero in locally flat regions and *I* is the identity matrix.

Our minimisation problem for PSF upsampling with dTV regularisation, under a non-negative constraint, is:

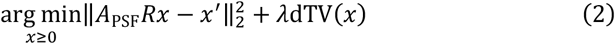

where *λ* is a regularisation parameter balancing the data fidelity and the regularisation.

The Primal-Dual Hybrid Gradient (PDHG) algorithm [31], provided by CIL [32] was used to solve (2). Here, the initial minimisation problem is decomposed into two subproblems, i.e., proximal operators, that have a closed-form solution. The proximal operator for dTV [26] is implemented using the fast gradient projection (FGP) algorithm [33] and provided by the CCPi-Regularisation Toolkit [34]. In addition, the dTV term enforces non-negativity by masking negative values in the reconstruction estimate at each FGP iteration.

### 2.2 Comparison with segmentation-guided partial volume correction

The PETPVC [35] library is an open source, commonly used library that implements a number of traditional PVC techniques for PET image. A literature search reveals that the RBV approach is most commonly applied in literature, however in this work we compare against the IY approach, as a logical extension of RBV:

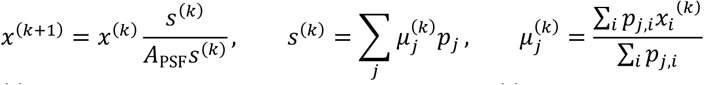

where *x*^(k)^ represents the corrected image at iteration *k, s*^(k)^ is the segmented constant image representing average values at each region, *j, μ*^(k)^ are the estimated regional means at iteration *k* for region *j*, and *p*j are the probability maps for each region, also indexable by voxel index, *i*.

Thomas *et al*. recognise that regional-based corrections fail to correct within regions, which is the case where an unsegmented lesion is present. Hence, we also compare with their proposed additional step of deconvolution. We investigated both IY + Richardson-Lucy (RL) and IY + reblurred Van-Cittert (rVC). For IY+RL, only one iteration was performed before images would become dominated by noise, whereas 10 iterations were performed for IY+rVC

### 2.3 Simulation study

Synthetic results were developed using the BrainWeb atlas [36], [37], which was used to provide template T1, FDG (4:1 grey to white matter contrast ratio) and probabilistic tissue map (CSF, grey matter, white matter) brain scans at 1×1×1 mm resolution. To avoid a piece-wise constant ground truth, each tissue class of each of the images had independent, low-frequency (0.5 mm^-1^) noise was added to each tissue [23]. An additional simulated pathological ground truth image set was generated in which simulated lesions were included in the PET image, with no matching contrast in the T1 image, in order to introduce unmatched features.

An analytic PET simulation was conducted to simulate the PET acquisition process using SIRF wrapping of STIR [38], [39]. PET images were resampled to 2.09×2.09×2.03 mm, the default reconstruction resolution on the Siemens Biograph mMR, and forward projected using a projector modelled on the mMR geometry that pre-blurs with a Gaussian filter of 4.5×4.5×4.5 mm FWHM [23] to simulate the scanner PSF and models detector size. The sinogram was then Poisson sampled to simulate an acquisition with 10^7^ counts. The effects of random coincidences, attenuation and scatter were not considered. The sinograms were subsequently reconstructed to 2.09×2.09×2.03 mm^3^ voxel-size using OSEM with 7 subsets and 40 iterations, before images were resampled to the 1×1×1 mm high resolution. No PSF model was included in the projectors during reconstruction to introduce an unmatched acquisition model.

### Determination of forward model

The forward model for the deconvolution problem is approximated as a Gaussian blurring kernel, (1). In order to determine appropriate parameters for the forward model, σXy and σz a simple, piece-wise constant digital phantom [40] was simulated with the analytic PET simulation described in Section 2.3. The optimal filter widths for the forward model were optimised using the Optuna hyperparameter optimization library [41] to minimise the outer optimisation:

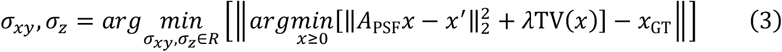

where *x*GT is the ground truth PET distribution. Here, TV was selected for regularisation to suppress noise in the deconvolution and *λ* was chosen such that regularisation was empirically low. As such, the deconvolution problem was minimally regularised to prevent it from biasing the optimisation of the forward model.

### 2.4 Application to a clinical 18F-florbetaben patient

A healthy volunteer with a high polygenic risk score for Alzheimer’s Disease was administered with 300 MBq of the amyloid tracer, 18F-florbetaben (Ethical approval by QIMR HREC P2193). After 90 mins of uptake, the participant was scanned for 20 minutes on a Siemens Biograph mMR. PET images were reconstructed to 2.09×2.09×2.03 mm^3^ voxel size. The CapAIBL analysis software [42] was used to generate a segmentation using an atlas-based approach.

#### Determination of forward model

As no ground truth is available, the method outlined in (3) is inappropriate. Instead, a manual optimisation was performed, again using an under-regularised TV prior, but relaxing the non-negativity requirement. σXy and σz were increased in increments of 0.5 mm and the last value before noticeable negative undershoot was seen outside the scalp.

## 3 Results

### 3.1 Simulation Study

**Forward model** optimisation found that the Gaussian forward model was optimally matched at σXy = 5.468 mm FWHM, σz = 5.269 mm FWHM, which was optimised with *λ* = 0.1. Fig. 1 shows the input, noisy and deconvolved spheres phantom, demonstrating contrast recovery with minimal blurring at edges. Residual blurring and noise are due to deliberate under-regularisation and edge blurring is approximately symmetric in the *x, y* and z axes. An accuracy of ±0.1 mm yielded virtually identical results, so σXy = 5.5 FWHM, σz = 5.3 FWHM were used for the BrainWeb experiments.

**Fig. 1.**
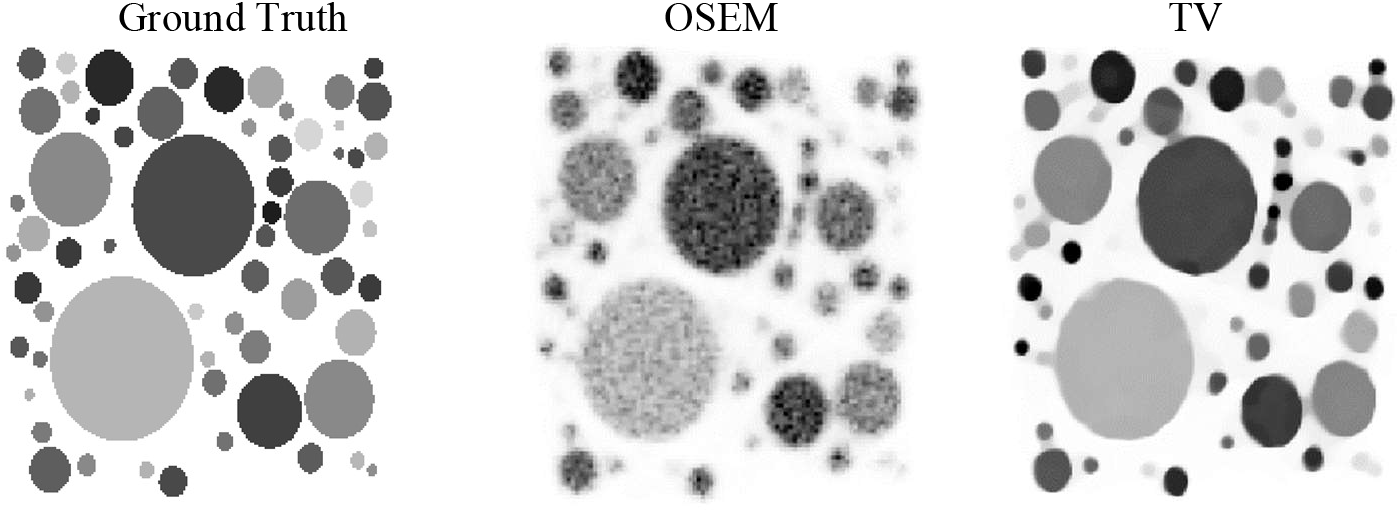
Results after optimising the Gaussian forward model to deconvolve the OSEM reconstruction (middle) to maximise similarity between the ground truth (left) and TV regularised deconvolution (right).

**Healthy simulation results** are depicted in Fig. 2. *λ* = 4 was empirically chosen for dTV, although the results were not observed to be highly dependent on the parameter. Summary statistics are presented in Table 1.

**Table 1.** Healthy BrainWeb simulation and deconvolution regional statistics.

|  | GM mean | GM SD | WM mean | WM SD | GM/WM ratio |
| --- | --- | --- | --- | --- | --- |
| Ground Truth | 120.878 | 14.938 | 35.611 | 9.179 | 3.394 |
| OSEM | 89.520 | 25.853 | 58.957 | 23.578 | 1.518 |
| IY | **115.674** | 28.746 | **39.221** | **12.628** | **2.949** |
| IY+RL | 105.107 | 56.303 | 42.992 | 24.852 | 2.445 |
| IY+rVC | 127.093 | 43.564 | 43.695 | 26.563 | 2.909 |
| dTV (proposed) | 111.556 | **20.786** | 42.950 | 15.876 | 2.597 |

**Fig. 2.**
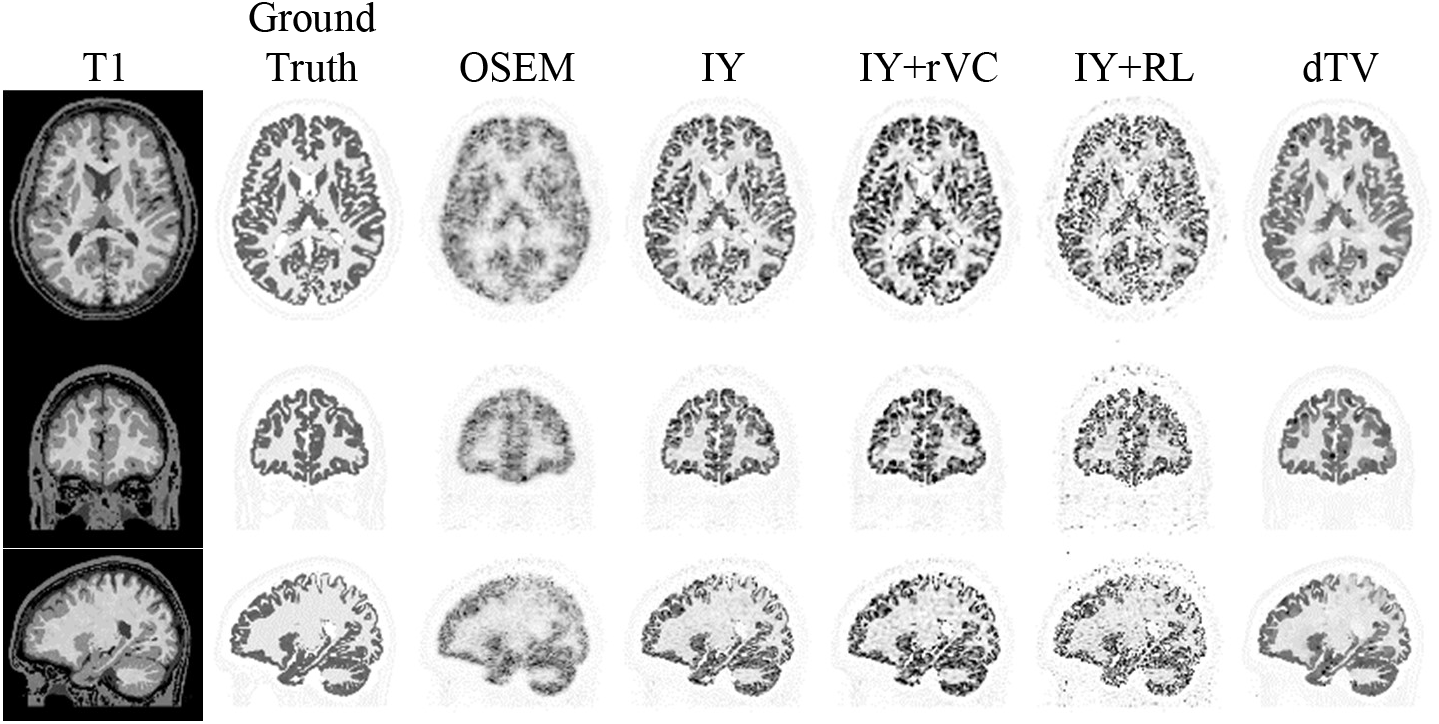
Healthy BrainWeb simulation and deconvolution. Axial, coronal and sagittal (top to bottom) views for the T1 anatomical prior, FDG ground truth, OSEM reconstruction, and PVC images using IY, IY+rVC, IY+RL and dTV.

**Pathological simulation results**, where PET-unique lesions were added to the ground truth, are illustrated in Fig. 3. Line profiles for Tumour 1 are given in Fig. 4.

**Fig. 3.**
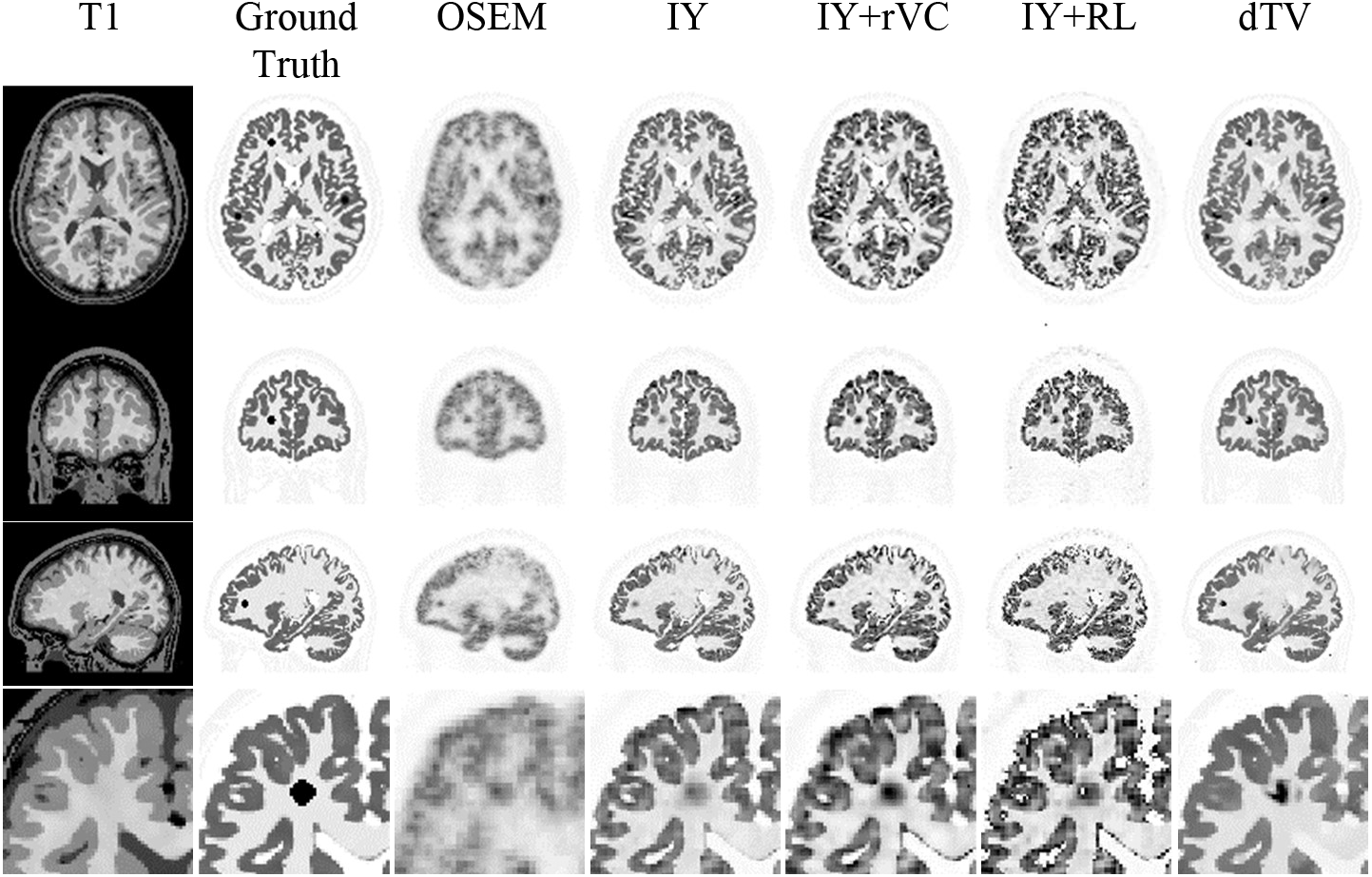
Pathological BrainWeb simulation and deconvolution. Layout is per Fig. 2, with an additional row showing an inset of Tumour 1.

**Fig. 4.**
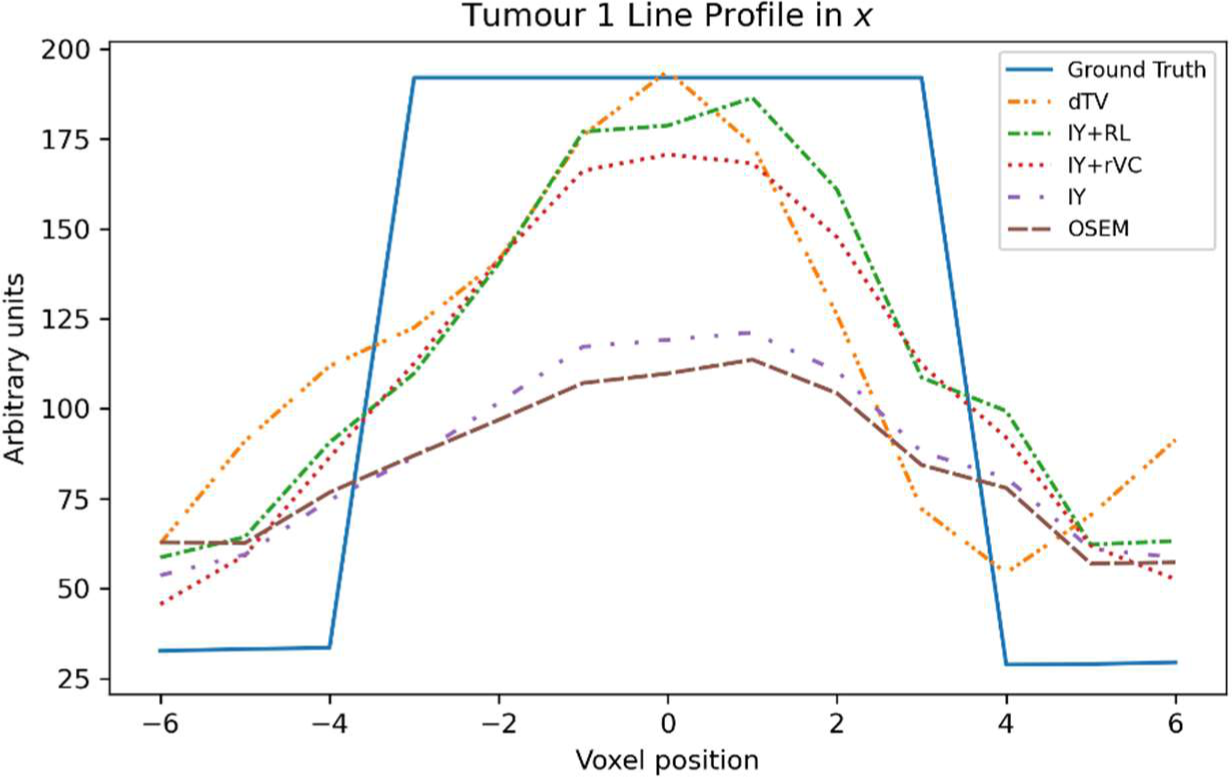
Line Profile of the simulated lesion in the x direction. Ground truth is compared against OSEM reconstruction and PVC images using IY, IY+RL, IY+rVC and dTV.

**Clinical patient results** are depicted in Fig 5. The optimal forward found was σXy = 4 mm FWHM and σz = 2 mm FWHM. The IY+RL results were omitted, as the algorithm failed to conserve activity and recovered the WM to unrealistic values. Summary statistics are presented in Table 2.

**Fig. 5.**
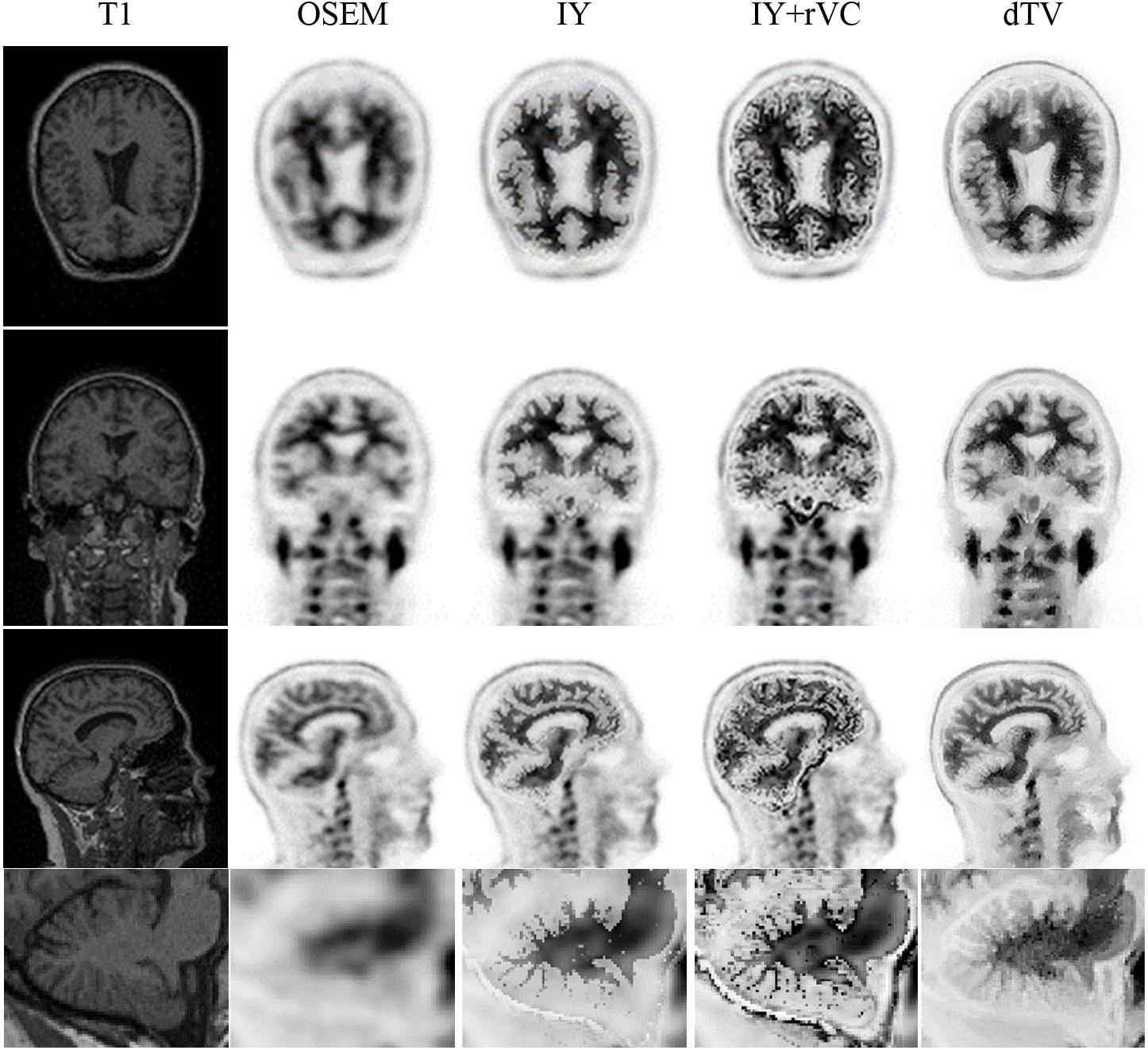
Pathological BrainWeb simulation and deconvolution. Layout is per Fig. 2, with an additional row showing an inset of cerebellum.

**Table 2.**
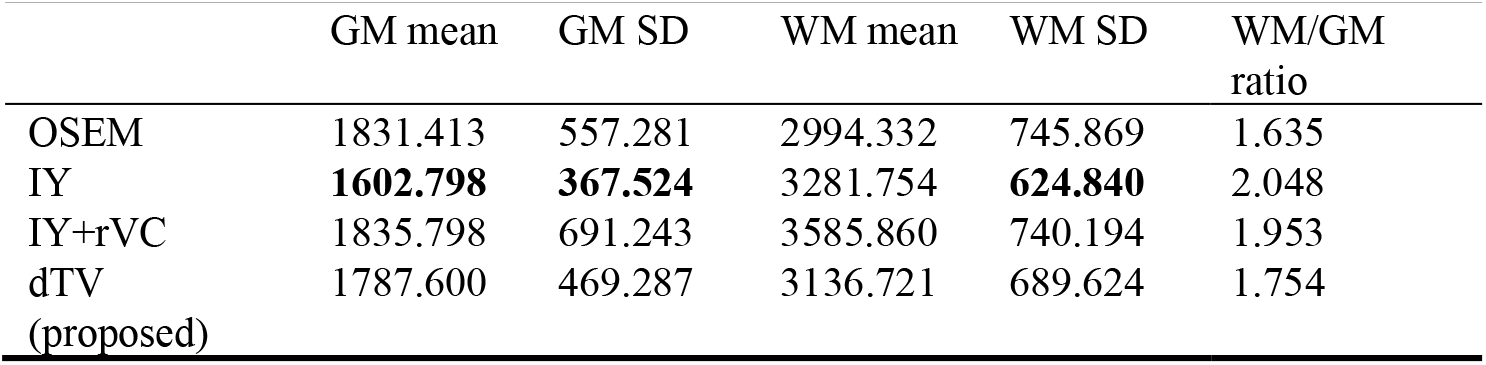
Clinical 18F-florbetaben and deconvolution regional statistics.

## 4 Discussion

### dTV PVC offers partial contrast recovery without ROI segmentation

All deconvolution methods were able to empirically improve the resolution in the cortex (Fig 2). All techniques had difficulty recovering the cortex in the superior parietal lobe, where the cortex is very thin. This was best recovered by IY (Fig 2, sagittal plane), which was also best able to recover gray/white matter (GM/WM) contrast (Table 1). However, the IY technique was unable to recover contrast in PET-unique lesions (Fig 3, 4). dTV was best able to recover contrast in the lesion, fully recovering the peak intensity, followed by IY+RL then IY+rVC. (Fig 4). The tumour recovery of the IY+RL technique came at the expense of increased noise, especially in the GM, such that the tumour was not the most intense feature in the lobe. This could likely lead to false positive detection.

### dTV PVC suppresses noise, but segmented PVC better preserves features

dTV was most successful in supressing noise (Fig 2, 3), also evident by the decreased GM SD (Table 1). Although noise in the WM is visually reduced (Fig 2, 3), the increased WM SD (Table 1) may be an indication that there is a bias in the dTV results near the WM/GM boundary. An advantage of the technique is that the *λ* parameter that allows a trade-off between noise suppression and model consistency. The sensitivity of this parameter was low and simple to optimise.

### dTV is robust in the presence of mis-segmentation

dTV was able to produce a satisfactory high resolution PET image in the cerebellum of the presented clinical patient (Fig. 5). The fine GM/WM boundary in this area proves a difficult segmentation problem, and the IY technique demonstrated poor results in regions where the segmentation was poor. IT+rVC was able to partially compensate.

### Limitations

Image space PSF approaches are inadequate for reliable and consistent high resolution PET imaging due to their inability to determine a specific PSF kernel for each crystal pair [43]. Sinogram space [7], [44], and hybrid space [43] PSF correction can overcome this limitation but cannot be implemented in the presented framework.

### Future Work

The current method uses a spatially invariant filter, which may not be optimal for all PET systems. Spatially variant filters, which can vary across the field of view, more accurately model the point spread function and improve image resolution [7] and can be implemented in image space [3]–[6]. Future work on this method will focus on incorporating spatially variant filters.

In clinical PET imaging, a true, high resolution validation is unavailable. Therefore, the most accurate validation of such techniques is in determining their contribution toward improving the effect size in a clinical study.

## 5 Conclusion

In this work, we have presented a preliminary demonstration of a dTV-PVC approach for PET super-resolution in combined PET/MR. dTV-PVC uses PHDG to optimise a deconvolution problem regularised by the dTV of the resolved PET with respect to an anatomical MR prior without segmentation. Compared with traditional segmentation-driven PET PVC approaches, the technique exhibits an attractive trade-off between tissue contrast recovery, PET-unique lesion recovery and noise suppression.

## Data Availability

All non-human data produced in the present study are available upon reasonable request to the authors

## Acknowledgements

The authors thank Kjell Erlandsson for providing advice on the comparison techniques implemented in PETPVC. The Prospective Imaging Study of Ageing investigators provided the clinical PET data. The CCP-SyneRBI and CCPi provided a forum for collaboration to facilitate this work. This work made use of computational support by CoSeC, the Computational Science Centre for Research Communities, through CCPi, and use of software infrastructure provided by the CCP-SyneRBI.

